# Research attitudes mediate the association between educational attainment and willingness to participate in Alzheimer’s disease research beyond Alzheimer’s disease knowledge

**DOI:** 10.64898/2026.10.13.26365095

**Authors:** Kyra M. Bonta, Emma Corbett, Meseret K. Alemu, Beniam D. Darge, Lauren K. Hilderbrand, Kaitlyn Richardson, Isabella M. Blum, Stephanie S. Ogonuwe, Kevin J. Shen, Madison K. McGuire, Diya H. Pandhi, Lewis W Gallagher, Jennifer M. Hunter, Fredrica Grant, Jackiethia Butsch, Neale Davis, Wy’Esha Simmons, Loletia Davis, Pamela Shields, Sue Cooper, Missy Rouse, Kimberly S. Wilson-Lawson, Carlotta Hughes-Culp, Mary Howard, Mertice Monroe, Matthew Coleman, De’leah Leath, Jimmy Henry, Pamela Gregory, Jeremy G. Grant

**Affiliations:** Department of Psychology, The Ohio State University, Columbus, OH, USA; Department of Psychology, University of Michigan, Ann Arbor, MI, USA; University of Maryland School of Graduate Studies, Baltimore, MD, USA; Center for Autism Spectrum Disorders, Nationwide Children’s Hospital, Columbus, OH, USA; Office of Health Equity, The Ohio State University Wexner Medical Center, Columbus, OH, USA; The James Cancer Hospital, The Ohio State University Wexner Medical Center, Columbus, OH, USA; Urban Aging Residents Coalition, Columbus, OH, USA; Central Ohio Chapter, Alzheimer’s Association, Dublin, OH, USA; African American Alzheimer’s & Wellness Association, Columbus, OH, USA; St. Paul’s AME Church, Columbus, OH, USA; Church of Christ of the Apostolic Faith, Columbus, OH, USA; Tabernacle Baptist Church, Columbus, OH, USA; Eta Nu Nu Chapter, Omega Psi Phi Fraternity, Columbus, OH, USA; American Heart Association, Columbus, OH, USA; African American Male Wellness Agency, Columbus, OH, USA

**Author notes:** Corresponding author: Jeremy G. Grant, Ph.D., The Ohio State University, College of Arts and Sciences, Department of Psychology, 1835 Neil Avenue, Columbus, OH 43210.

**Keywords:** Research attitudes, recruitment, education, socioeconomic status, Alzheimer’s disease, dementia

## Abstract

**INTRODUCTION:** This study examined the extent to which educational attainment, Alzheimer’s disease (AD) knowledge, and research attitudes are associated with willingness to participate in AD research, and whether AD knowledge and research attitudes mediate associations between educational attainment and willingness to participate.

**METHODS:** 201 older adults completed self-report measures of AD knowledge, research attitudes, prior research participation, and willingness to participate in diagnostic procedures (neuropsychological assessment, blood draw, brain MRI) and non-pharmacological intervention research (tracking physical activity, aerobic exercise).

**RESULTS:** Favorable research attitudes were consistently associated with greater willingness to participate across all proposed research activities, and research attitudes mediated the relationship between educational attainment and willingness to participate in research. In contrast, AD knowledge did not mediate the association between educational attainment and willingness to participate.

**DISCUSSION:** Research attitudes may be a key, modifiable determinant of willingness to participate in AD research and a promising focus for broadening AD research participation.

## 1. INTRODUCTION

Alzheimer’s disease and Alzheimer’s disease-related disorders (AD/ADRD) disproportionately affect vulnerable populations, including individuals with low educational attainment, residents of socioeconomically disadvantaged and/or rural communities, and Black and Hispanic/Latino/a Americans (1–6). In addition to bearing greater disease burden, these populations remain underrepresented in AD research, limiting the generalizability of findings and contributing to widening disparities in dementia prevention, diagnosis, and treatment (1,2,7–10). Consequently, concerns remain regarding the equitable translation of advances in dementia research to populations that have been insufficiently represented in AD research (1,2,9,10).

Importantly, underrepresentation in AD research does not necessarily reflect a lack of willingness to participate among these populations (11,12). Evidence suggests that Hispanic and Non-Hispanic Black Americans are just as willing as non-Hispanic White Americans to participate in health research in general, but often report fewer opportunities to do so (12,13). Within AD research, several studies have demonstrated high levels of willingness among Black older adults and individuals with lower educational attainment and socioeconomic status (14). Similarly, individuals residing in socioeconomically disadvantaged neighborhoods have reported willingness to participate in AD research comparable to that of individuals living in more advantaged neighborhoods, despite being underrepresented in recruitment registries (15). These findings suggest that disparities in research participation may be driven less by unwillingness and more by factors such as inadequate outreach by research organizations, logistical burdens, and other structural barriers to enrollment. Therefore, there is a pressing need for evidence-based approaches to bolstering the representativeness of AD research (1,8,9). Identifying factors associated with willingness to participate in AD research may help inform the development of more effective recruitment strategies.

An individual’s willingness to participate in AD research may be influenced by their attitudes toward biomedical research and its perceived benefits. Previous studies have linked favorable research attitudes to greater willingness to participate in research and improved retention in research registries (16). Research attitudes also appear to influence engagement beyond initial enrollment. In a study of participants enrolled in an AD clinical trial, Stites et al. found that individuals with more favorable baseline research attitudes were significantly more likely to complete the trial and adhere to study procedures than those with less favorable attitudes (17). Collectively, these findings suggest that research attitudes may represent a modifiable target for interventions aimed at bolstering the recruitment of representative research samples.

More research is needed to understand how research attitudes interact with other factors associated with willingness to participate in AD research, including educational attainment (18,19). and AD knowledge (20,21). Higher educational attainment has been associated with greater engagement in AD research, including higher rates of study completion, retention, and enrollment (18,19). Given that educational attainment may facilitate the acquisition and application of health information, AD knowledge may represent one mechanism through which education influences research participation. Educational interventions designed to improve AD knowledge have been shown to increase research attitudes and subsequent enrollment in studies (20). Consistent with these findings, the Knowledge, Attitudes, and Practices (KAP) framework suggests that increased knowledge about a particular topic can foster positive attitudes, which can subsequently influence decision-making and behavior (22). Supporting this framework, one study found a trend toward greater willingness to undergo a dementia screening among individuals with higher dementia literacy (21). However, attitudes can also be disconnected from knowledge and behavior. For example, a recent cross-sectional study of visitors to a regional hospital in China found that middle-aged and older adults expressed favorable attitudes toward dementia risk-reduction activities despite reporting limited knowledge of modifiable dementia risk factors and low participation in such behaviors (23).

Although educational attainment, AD knowledge, and research attitudes have each been linked to research engagement, few studies have examined the simultaneous contributions of these characteristics within a single analytic framework. Accordingly, the present study aimed to (1) examine the extent to which educational attainment, AD knowledge, and research attitudes are associated with willingness to participate in AD research procedures and (2) determine whether AD knowledge and research attitudes mediate the associations between educational attainment and willingness to participate in a community-based sample of predominantly Black older adults. We hypothesized that higher educational attainment, AD knowledge, and research attitudes would be associated with greater willingness to participate in three diagnostic procedures (undergoing a neuropsychological assessment, undergoing a blood draw to detect blood-based biomarkers of AD, and undergoing a brain MRI) and two research activities involving non-pharmacological interventions to promote brain health (tracking physical activity with a smartwatch and engaging in aerobic exercise). We also hypothesized that higher educational attainment would be associated with greater AD knowledge and more positive research attitudes, which would sequentially predict a greater willingness to participate in all five research activities.

## 2. METHODS

### 2.1. Participants

Participants were 201 community-dwelling adults residing in central Ohio who were between 50 and 89 years of age and fluent English speakers. Individuals were excluded if they had visual or auditory impairments that would interfere with questionnaire completion, a known diagnosis of dementia, or were otherwise unable to provide informed consent independently.

Participants were recruited through various community engagement initiatives in Central Ohio, including tabling events at health fairs, brain health presentations delivered in collaboration with local faith communities and community organizations, and health screening events conducted in partnership with clinics serving underserved populations. A community advisory board consisting of representatives from national (Alzheimer’s Association, American Heart Association, African American Male Wellness Agency) and local organizations (African American Alzheimer’s and Wellness Association, Urban Aging Residents Coalition) provided feedback on the questionnaires and partnered with the research team to recruit participants. All participants received a $10.00 gift card for completing the self-report questionnaires.

### 2.2. Measures

#### 2.2.1. Research Attitudes Questionnaire (RAQ)

The Research Attitudes Questionnaire (RAQ) is a brief self-report measure consisting of seven items that assess attitudes toward biomedical research (24). Items reflect perspectives on the societal value of research, trust in researchers, perceived safety of research participation, optimism about research outcomes, and altruistic views regarding volunteering for research. Responses are rated on a 5-point Likert scale ranging from 1 (strongly disagree) to 5 (strongly agree). Item responses are summed to produce a total score ranging from 7 to 35, with higher scores indicating more favorable attitudes toward biomedical research (24).

#### 2.2.2. Willingness to Participate in Research

Willingness to participate in research was assessed using five items related to specific diagnostic procedures and behavioral interventions available to the research team in the Department of Psychology at The Ohio State University. Participants indicated how willing they would be to (1) undergo a neuropsychological assessment, (b) undergo a blood draw to assess blood-based biomarkers of AD, (c) undergo a brain MRI, (d) use a smartwatch to track physical activity, and (e) engage in moderate-intensity aerobic exercise. Responses were rated on a 4-point Likert scale: not interested, somewhat interested, moderately interested, and very interested.

#### 2.2.3. AD Knowledge (BKAD)

AD knowledge was assessed using the Basic Knowledge of Alzheimer’s Disease (BKAD) questionnaire, a 32-item measure designed to evaluate general understanding of Alzheimer’s disease, including common symptoms and disease trajectories (25). The questionnaire includes statements addressing topics such as risk factors, early symptoms, treatment and management, and lifestyle factors associated with dementia risk. Participants indicate whether each statement is true or false, and responses are scored as correct or incorrect. Correct responses are summed to yield a total score ranging from 0 to 32; higher scores reflect greater knowledge of Alzheimer’s disease.

#### 2.2.4. Neighborhood Socioeconomic Status

The Area Deprivation Index (ADI) is a validated measure of neighborhood SES based on 17 indicators of income, education, employment, and housing quality (26). ADI values are generated at the census block group level and can be reported using either national percentile rankings or state-specific rankings. In the present study, ADI state deciles were used, with higher values indicating greater neighborhood disadvantage relative to other areas within the same state. Participants’ 9-digit ZIP codes were used to assign the corresponding census block group ADI value. For analysis, state ADI deciles were collapsed into five categories (deciles 1–2, 3–4, 5–6, 7–8, and 9–10), with higher categories indicating greater neighborhood socioeconomic disadvantage.

#### 2.2.5. Previous Research Participation, Family History of Dementia, and Demographic Characteristics

Participants responded using dichotomous (yes/no) response options regarding family history of dementia and previous participation in research at Ohio State University. They also provided demographic information, including age, race, education, employment status, income, transportation access, health insurance status, and the presence of any cardiovascular risk factors.

### 2.3. Procedures

All study procedures complied with the Declaration of Helsinki. The study protocol was reviewed by The Ohio State University Biomedical Sciences Institutional Review Board and was granted exempt status. Data were cross-sectional and collected at a single time point. Participants completed a series of questionnaires either in person (*n* = 161) or online (*n* = 40), including the RAQ, BKAD, and customized questions assessing willingness to participate in future research. For in-person participation, participants underwent verbal consent and completed the self-report questionnaires independently. Participants completing the study remotely provided informed consent electronically and completed self-report questionnaires via a secure REDCap portal.

### 2.4. Statistical Analyses

Statistical analyses were conducted using SPSS v.31.0. Five ordinal logistic regression models were run, with willingness to undergo a neuropsychological assessment, undergo a blood draw, undergo an MRI, use a smartwatch to track physical activity, and engage in aerobic exercise as outcome variables. Predictor variables included age, years of education, BKAD total score, RAQ total score, family history of dementia (a proxy for the personal relevance of dementia research), and prior research participation at The Ohio State University. Due to small cell sizes in the “not interested” response category across the outcome variables (3.5%–14.4% of participants across the five research participation variables), each outcome variable was recoded into three levels. Responses of “not interested” and “somewhat interested” were combined to form a low-interest category, while “moderately interested” and “very interested” were retained as separate categories.

Serial mediation analyses were conducted using PROCESS version 4.2 for SPSS (Model 6) to examine whether the association between years of education and willingness to participate in research was mediated by BKAD total score and RAQ total score. The models included education as the predictor, BKAD total score as the first mediator, RAQ total score as the second mediator, and each of the five willingness-to-participate variables as the outcome. Age was included as a covariate. Indirect effects were estimated using 5,000 bootstrap samples, and effects were considered statistically significant when the 95% bootstrap confidence interval did not include zero. Two participants with RAQ scores of 7, the minimum possible score on the scale, were identified as influential outliers and excluded from the primary analyses; the next-lowest observed RAQ score was 14. Sensitivity analyses including these two participants produced the same pattern of results reported below.

## 3. RESULTS

### 3.1. Participant Characteristics

Table 1 displays the descriptive characteristics of the overall sample. The final sample included 201 community-dwelling adults, with a mean age of 65.5 ± 9.5 years. The sample was predominantly female (81.6%) and Black (79.6%). Other demographic variables were generally comparable to the demographics of central Ohio. Overall, participants appeared to be relatively well resourced, with moderate levels of educational attainment and household income, high rates of health insurance coverage, and reliable access to transportation. Participants were distributed across all Area Deprivation Index (ADI) state deciles, although most resided in lower-deprivation areas. Of the various recruitment modalities, the largest proportion of participants were recruited through health fairs (41.8%).

**Table 1.** Demographic characteristics for the overall sample (*n* = 201)

| **Variable** | **All Participants** |
| --- | --- |
| Age (years) | 65.5 ± 9.5 |
| Age group |  |
| 50-59 | 30.3% |
| 60-69 | 34.8% |
| 70-79 | 26.9% |
| 80-89 | 8.0% |
| Gender (% women) | 81.6% |
| Primary Language |  |
| English | 89.6% |
| Spanish | 0.5% |
| Other | 1.5% |
| Race / ethnicity |  |
| American Indian/Alaska Native | 0.5% |
| Asian | 1.5% |
| Black (non-Hispanic) | 79.6% |
| Hispanic/Latino/a | 0.5% |
| White (non-Hispanic) | 13.4% |
| Mixed Race | 4.5% |
| Education level |  |
| Less than high school | 0.5% |
| High School Diploma/GED | 13.4% |
| Some college/ Associate's degree | 24.9% |
| Bachelor's Degree | 30.8% |
| Master's degree | 25.4% |
| Professional/Doctoral Degree | 5.0% |
| Employment Status |  |
| Employed | 39.3% |
| Retired | 54.2% |
| On Disability | 4.0% |
| Unemployed | 1.0% |
| Prefer not to Answer | 1.5% |
| Income level |  |
| \$0 - \$29,999 | 16.9% |
| \$30,000 - \$74,999 | 34.8% |
| \$75,000 and over | 36.3% |
| Unknown Income | 11.9% |
| Reliable access to transportation (% yes) | 95.5% |
| Access to healthcare insurance (% yes) | 46.3% |
| ADI - State Decile |  |
| 1-2 | 31.3% |
| 3-4 | 28.8% |
| 5-6 | 17.4% |
| 7-8 | 11.4% |
| 9-10 | 10.9% |
| Number of cardiovascular risk factors |  |
| 0 | 22.4% |
| 1 | 37.8% |
| 2 | 27.4% |
| 3 | 9.0% |
| 4 | 3.5% |
| What type of event was this participant recruited from? |  |
| In-Person Presentation | 24.9% |
| Health Fair | 41.8% |
| Other tabling event | 13.4% |
| Online | 19.9% |

**Note.** ADI = The Area Deprivation Index. The number of cardiovascular risk factors was calculated using the sum of the number of health conditions; high blood pressure, high cholesterol, high blood sugar/diabetes, and heart disease.

Figures 1.1 and 1.2 illustrate participants’ willingness to engage in AD-related research activities and interventions. Across all activities, the largest proportion of participants reported high willingness, particularly for completing a neuropsychological assessment, undergoing a blood draw, participating in smartwatch-based physical activity tracking, and engaging in aerobic exercise. Willingness to undergo a brain MRI was more evenly distributed across low- and high-willingness categories, although high willingness remained the most frequently reported response.

**Figure 1.1.**
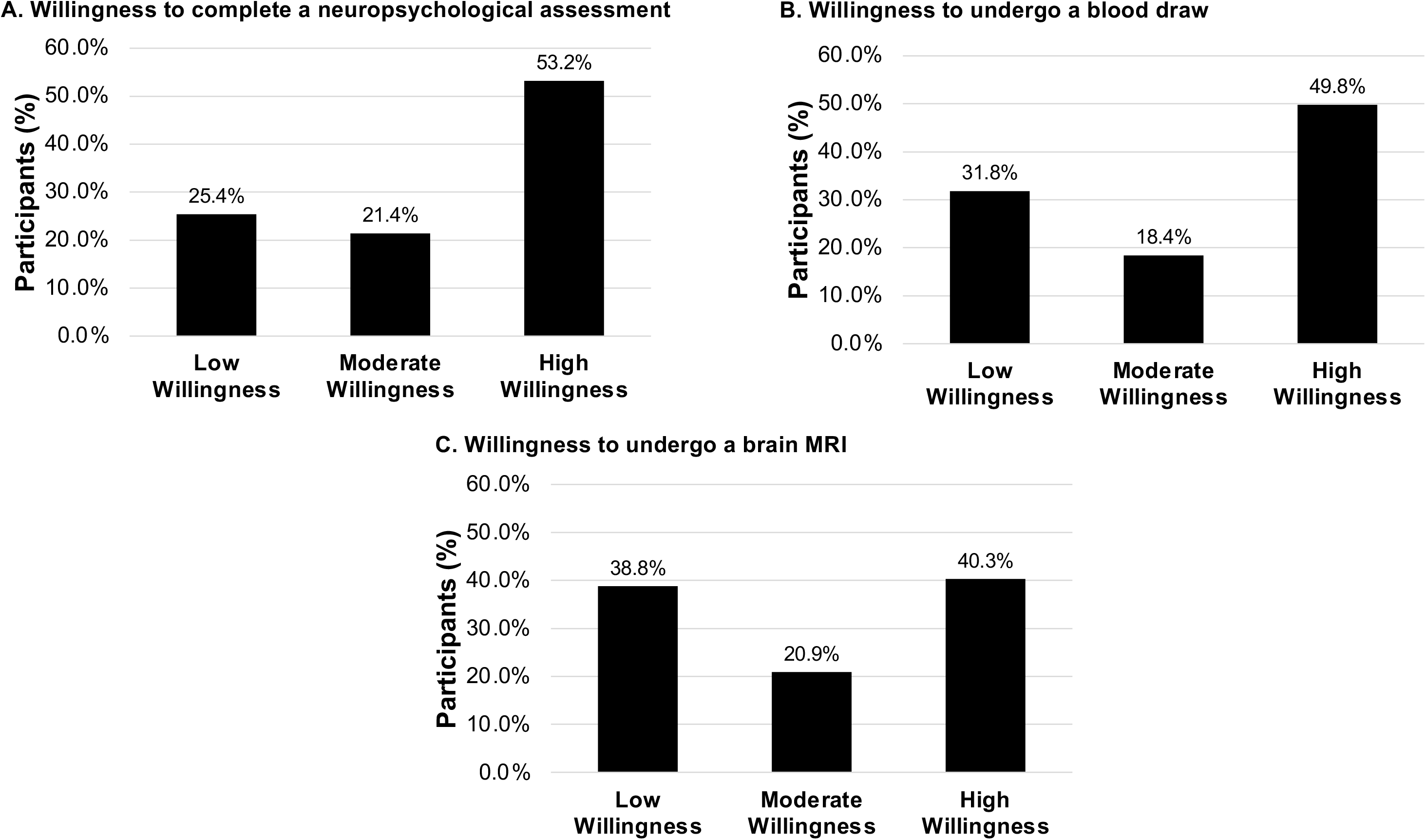
Distribution of study participants across willingness categories for research activities related to AD diagnosis. **Note.** Bars represent the percentage of participants in each willingness category (low, moderate, and high willingness) for (A) completing a neuropsychological assessment, (B) undergoing a blood draw, and (C) undergoing a brain MRI.

**Figure 1.2.**
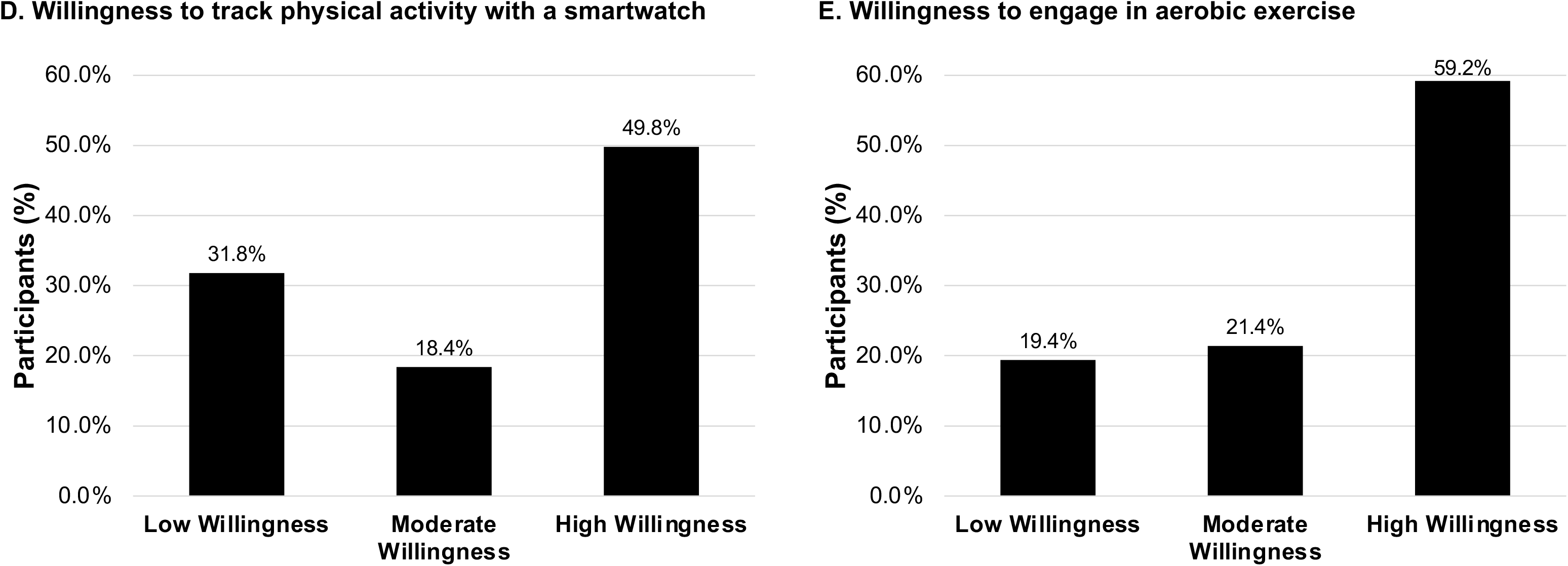
Distribution of study participants across willingness categories for research activities related to AD interventions. **Note.** Bars represent the percentage of participants in each willingness category (low, moderate, and high willingness) for (D) tracking physical activity using a smartwatch and (E) engaging in aerobic exercise.

**Figure 2.**
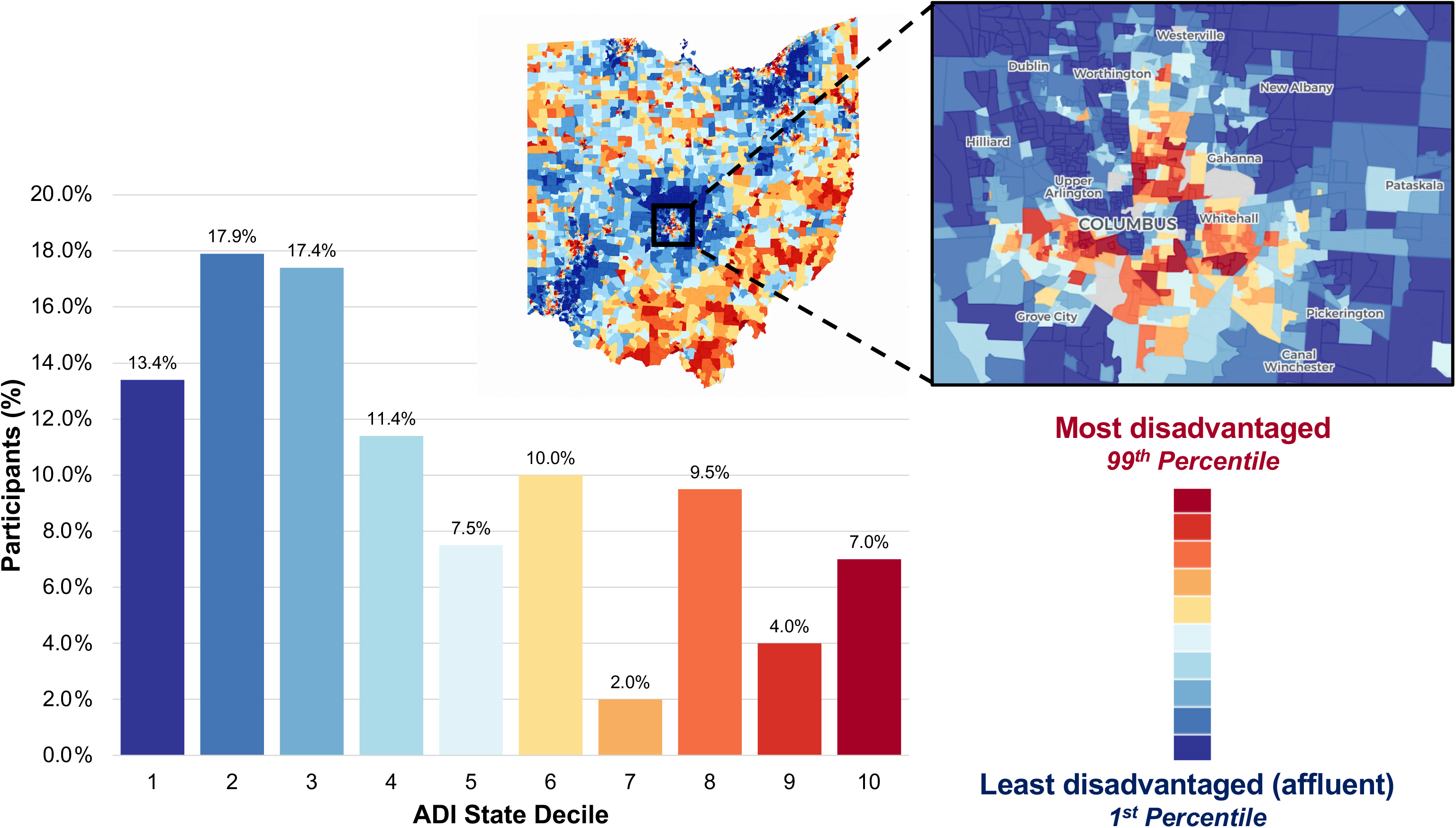
Distribution of study participants across State Area Deprivation Index (ADI) deciles and geographic variation in neighborhood disadvantage across Ohio. **Note.** Bars represent the percentage of participants within each State ADI decile. Higher ADI deciles indicate greater neighborhood socioeconomic disadvantage. The accompanying maps illustrate the geographic distribution of State ADI across Ohio census block groups and a magnified view of neighborhood-level variation within the Columbus metropolitan area. Blue indicates lower neighborhood disadvantage (1st percentile), whereas red indicates higher neighborhood disadvantage (99th percentile).

### 3.2. Participant Characteristics Across ADI State Categories

Table 2 presents participant characteristics, BKAD scores, RAQ scores, and willingness to participate in research activities across ADI state quintiles. Overall, participant profiles were generally similar across quintiles. Participants residing in the least disadvantaged neighborhoods (ADI deciles 1–2) tended to report slightly higher levels of education (15.9 ± 2.2 years), RAQ scores (27.3 ± 4.0), and BKAD scores (27.0 ± 4.2) than those residing in the most disadvantaged neighborhoods (ADI deciles 9–10; education: 14.6 ± 2.7 years, RAQ: 25.0 ± 4.5, BKAD: 25.6 ± 3.7). Notably, despite differences in neighborhood-level socioeconomic status, willingness to participate in research activities was broadly comparable across ADI quintiles. However, participants residing in neighborhoods in the middle ADI quintile (State ADI 5–6) tended to report lower willingness across several research activities, particularly those involving blood draws, neuropsychological assessment, brain MRI, and aerobic exercise.

**Table 2.** Demographic characteristics, AD knowledge, research attitudes, and willingness to participate in AD research activities by state ADI group.

| **State ADI Group** |  |  |  |  |  |
| --- | --- | --- | --- | --- | --- |
| **Variable** | **1-2**<br>**(n = 63)** | **3-4**<br>**(n = 58)** | **5-6**<br>**(n = 35)** | **7-8**<br>**(n = 23)** | **9-10**<br>**(n = 22)** |
| *Age (years)* | 65.8 ± 9.5 | 65.4 ± 9.0 | 65.7 ± 10.6 | 64.0 ± 8.7 | 66.0 ± 10.7 |
| *50-59* | 28.6% | 29.3% | 28.6 | 39.1 | 31.8 |
| *60-69* | 36.5% | 37.9% | 31.4 | 34.8 | 27.3 |
| *70-79* | 27.0% | 24.1% | 28.6 | 26.1 | 31.8 |
| *80-89* | 7.9% | 8.6% | 11.4 | - | 9.1 |
| *Years of Education* | 16.0 ± 2.2 | 16.0 ± 2.3 | 15.1 ± 2.2 | 15.1 ± 2.2 | 14.59 ± 2.7 |
| *BKAD Total Score*<br>*(range: 0-32)* | 27.0 ± 4.2 | 27.6 ± 3.8 | 25.9 ± 5.2 | 27.0 ± 4.4 | 25.6 ± 4.7 |
| *RAQ Total Score*<br>*(range: 7-35)* | 27.3 ± 4.0 | 27.3 ± 4.3 | 25.9 ± 3.7 | 25.5 ± 5.2 | 25.0 ± 4.5 |
| Willingness to undergo a neuropsychological assessment |  |  |  |  |  |
| *High Interest* | 57.1% | 56.9% | 42.9% | 52.2% | 50.0% |
| *Moderate Interest* | 20.6% | 13.8% | 34.3% | 17.4% | 27.3% |
| *Low Interest* | 22.2% | 29.3% | 22.9% | 30.4% | 22.7% |
| Willingness to undergo a blood draw |  |  |  |  |  |
| *High Interest* | 54.0% | 58.6% | 22.9% | 43.5% | 63.6% |
| *Moderate Interest* | 19.0% | 17.2% | 34.3% | 4.3% | 9.1.3% |
| *Low Interest* | 27.0% | 24.1% | 42.9% | 52.2% | 27.3% |
| Willingness to undergo a brain MRI |  |  |  |  |  |
| *High Interest* | 42.9% | 51.7% | 25.7% | 21.7% | 40.9% |
| *Moderate Interest* | 22.2% | 17.2% | 28.6% | 21.7% | 13.6% |
| *Low Interest* | 34.9% | 31.0% | 45.7% | 56.5% | 45.5% |
| Willingness to undergo track physical activity using a smartwatch |  |  |  |  |  |
| *High Interest* | 58.7% | 43.1% | 51.4% | 34.8% | 54.5% |
| *Moderate Interest* | 15.9% | 22.4% | 20.0% | 21.7% | 9.1% |
| *Low Interest* | 25.4% | 34.5% | 28.6% | 43.5% | 36.4% |
| Willingness to engage in moderate-intensity aerobic exercise |  |  |  |  |  |
| *High Interest* | 69.8% | 60.3% | 45.7% | 56.5% | 50.0% |
| *Moderate Interest* | 15.9% | 15.5 | 37.1% | 21.7% | 27.3% |
| *Low Interest* | 14.3% | 24.1% | 17.1% | 21.7% | 22.7% |

**Note:** RAQ = Research Attitudes Questionnaire (range: 7–35); BKAD = Basic Knowledge of Alzheimer's Disease (range: 0–32); ADI = Area Deprivation Index.

### 3.3. Ordinal logistic regressions predicting willingness to participate in AD research

Five ordinal logistic regression models were conducted to examine predictors of willingness to engage in AD research activities, including completing a neuropsychological assessment, a blood draw, a brain MRI, tracking physical activity using a smartwatch, and engaging in aerobic exercise. Regarding the three diagnostic procedures, participants generally expressed high willingness, although willingness tended to decrease as procedures became more burdensome. The proportion of participants who reported high willingness was greatest for the neuropsychological assessment (53.2%), somewhat lower for the blood draw (49.8%), and lowest for MRI (40.3%). Conversely, the proportion reporting low willingness increased from 25.4% for the neuropsychological assessment to 31.8% for the blood draw and 38.8% for undergoing a brain MRI. All five ordinal logistic regression models were statistically significant and met the proportional odds assumption (see Supplementary Tables S1-S5).

Across all models, the RAQ total score emerged as the most consistent predictor of willingness to participate in AD research. Higher RAQ scores were associated with greater willingness to complete a neuropsychological assessment (*B* = 0.118, *p* = .002), undergo a blood draw (*B* = 0.114, *p* = .002), undergo an MRI (*B* = 0.084, *p* = .016), participate in smartwatch-based exercise tracking (*B* = 0.090, *p* = .015), and engage in aerobic exercise (*B* = 0.111, *p* = .004).

Previous research participation was associated with greater willingness to complete a neuropsychological assessment (*B* = 0.852, *p* = .028) and undergo a blood draw (*B* = 0.831, *p* = .029). Younger age was associated with greater willingness to participate in smartwatch-based exercise tracking (*B* = -0.051, *p* < .001) and aerobic exercise (*B* = -0.039, *p* = .011). Years of education, family history of dementia, and BKAD total score were not significant predictors in any model (see Figure 3 and Supplementary Tables S1-S5).

**Figure 3.**
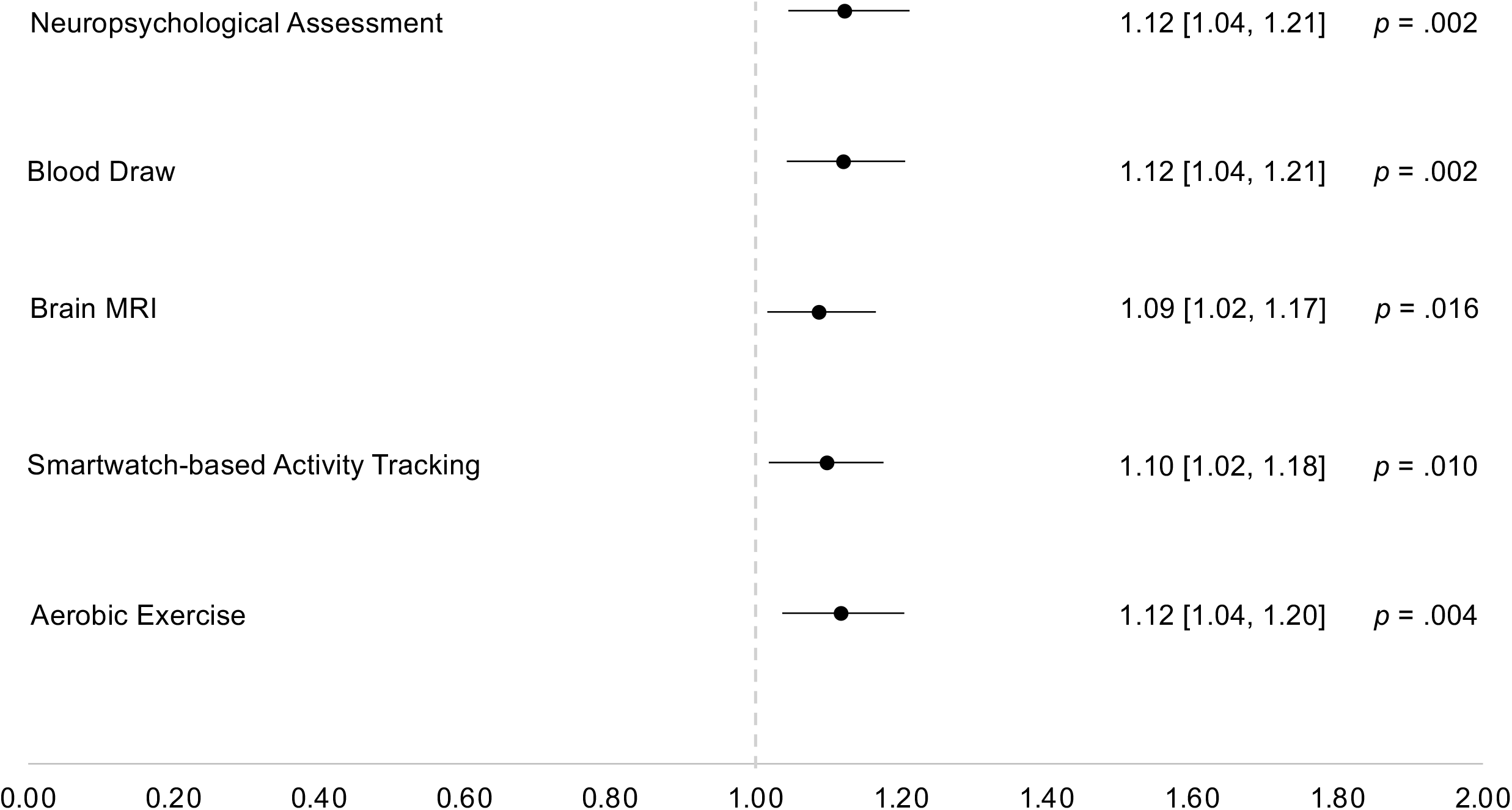
Odds ratios (95% CIs) for the association between research attitude scores and willingness to participate in Alzheimer’s disease (AD) research activities. **Note.** Odds ratios and 95% confidence intervals were derived from ordinal regression models examining the association between research attitude scores and willingness to participate in AD research activities. Odds ratios represent the change in the odds of greater willingness to participate associated with a one-unit increase in research attitude score. Results for all predictors included in the models are reported in Supplementary Tables 1–5.

### 3.4. Serial mediations predicting willingness to participate in AD research

Five serial mediation analyses were conducted to examine whether BKAD and RAQ scores sequentially mediated the association between years of education and willingness to participate in each of the five AD research activities (Figure 4). Age was included as a covariate in all models. Across all five models, greater years of education were associated with higher BKAD scores (*B* = 0.610, *p* < .001), and higher BKAD scores were associated with higher RAQ scores (*B* = 0.288, *p* < .001). In turn, higher RAQ scores were associated with greater willingness to participate in AD research activities (see Figures 5.1 and 5.2).

**Figure 4.**
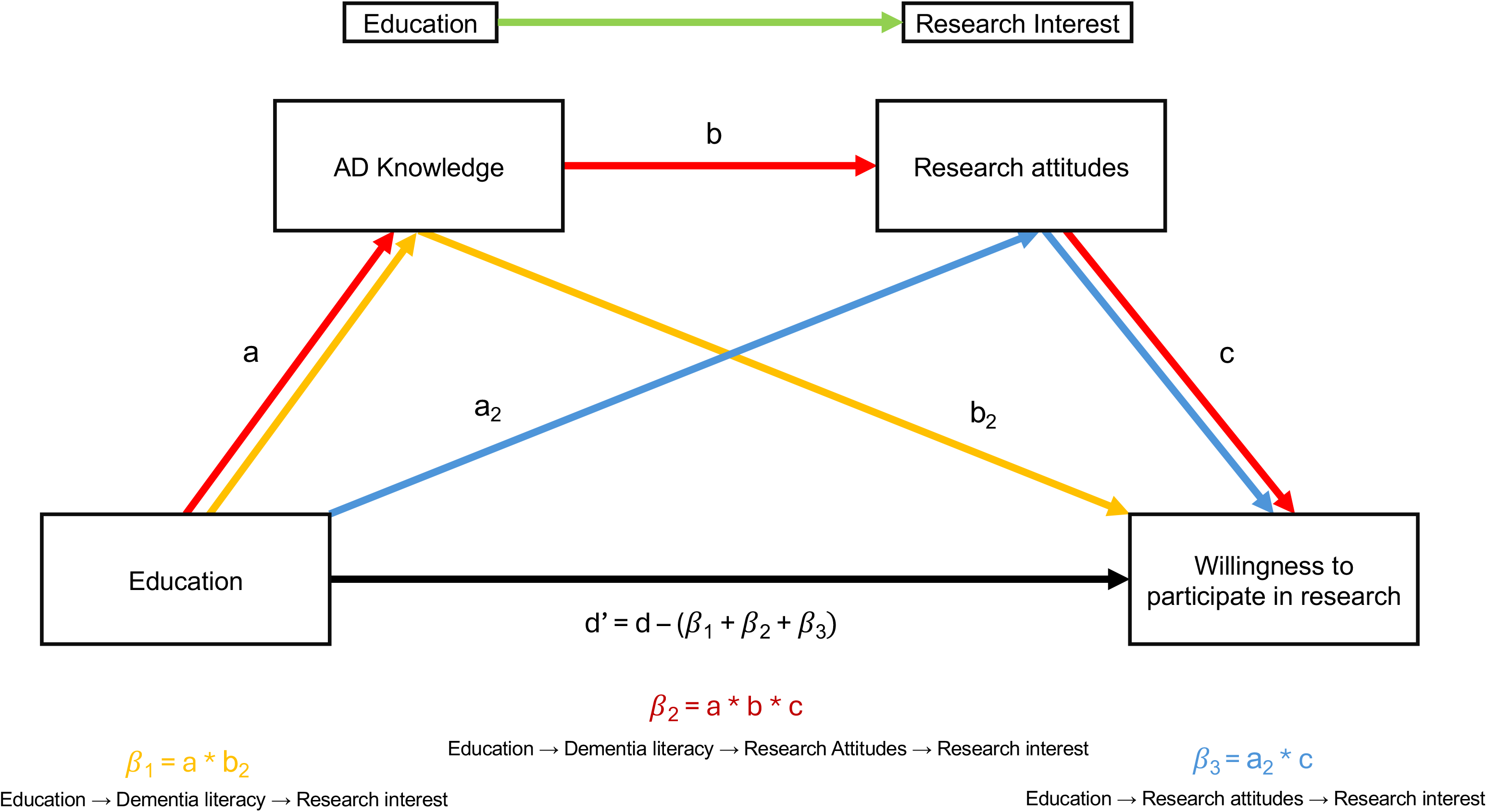
Conceptual serial mediation model illustrating the association between educational attainment and willingness to participate in AD research, with AD knowledge as the first mediator and research attitudes as the second mediator. **Note.** Colored pathways represent the three indirect effects evaluated in the serial mediation analyses: (β₁) education → AD knowledge → willingness to participate in research; (β₂) education → AD knowledge → research attitudes → willingness to participate in research; and (β₃) education → research attitudes → willingness to participate in research. The direct effect of educational attainment on willingness to participate in research is represented by d′.

**Figure 5.1.**
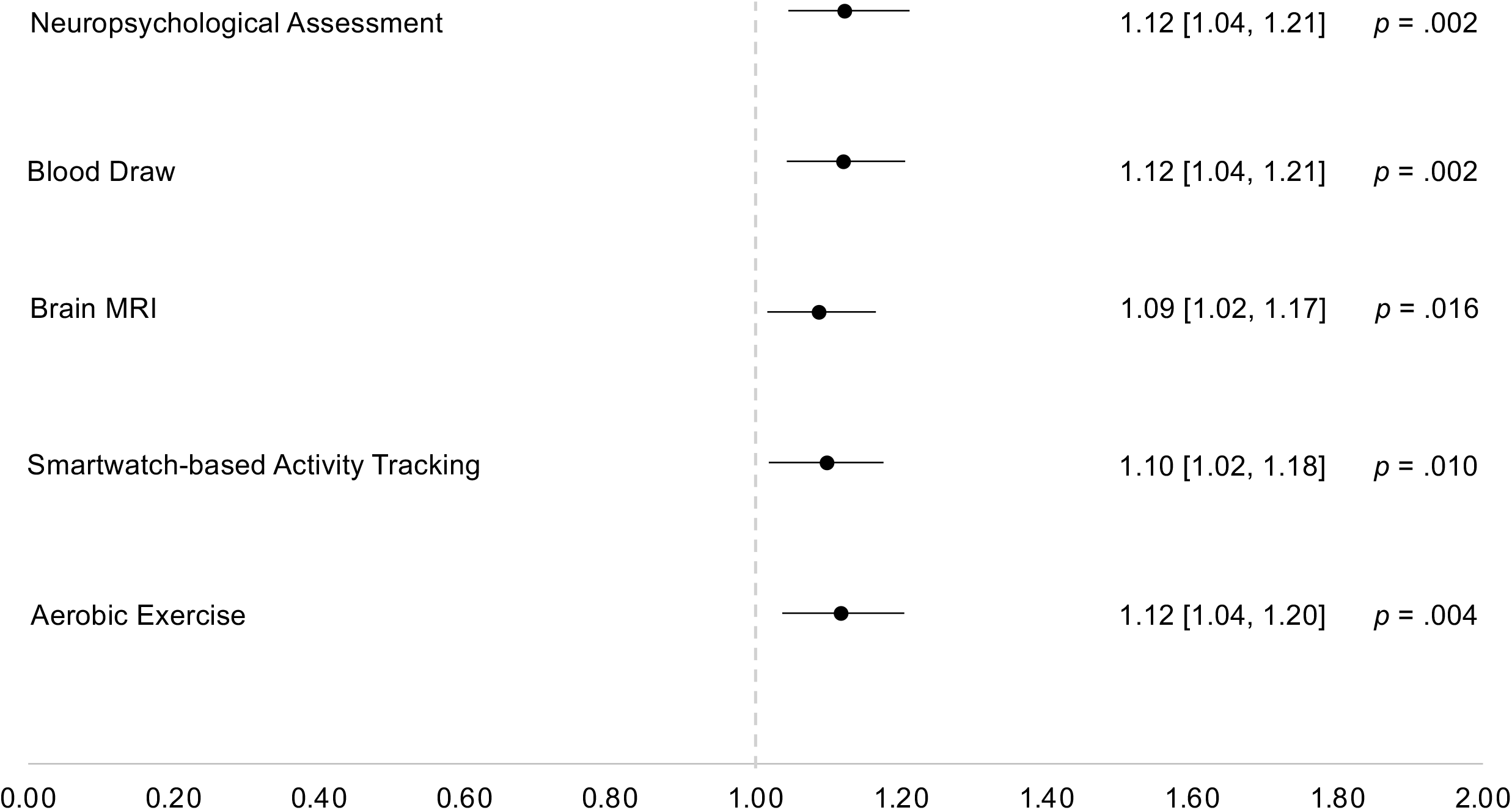
Serial mediation models examining the association between educational attainment and willingness to participate in research activities related to AD diagnosis, with AD knowledge as the first mediator and research attitudes as the second mediator. **Note.** Path coefficients for all modeled relationships are shown. Colored pathways indicate statistically significant indirect effects identified in the serial mediation analyses, whereas black pathways indicate non-significant relationships. Significant indirect effects and corresponding 95% confidence intervals are reported for each model.

**Figure 5.2.**
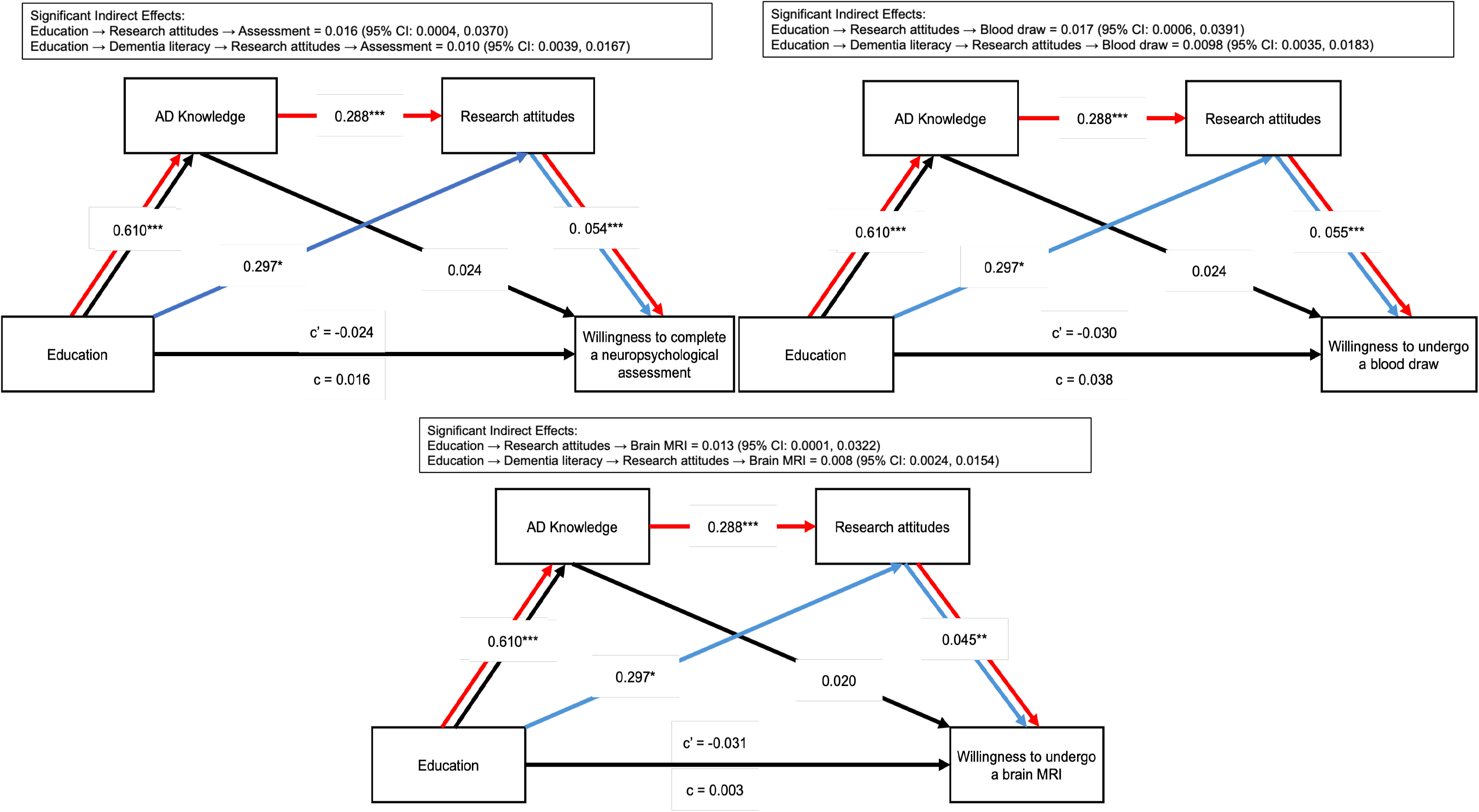
Serial mediation models examining the relationship between educational attainment and willingness to participate in research activities related to AD interventions, with AD knowledge as the first mediator and research attitudes as the second mediator **Note.** Path coefficients for all modeled relationships are shown. Colored pathways indicate statistically significant indirect effects identified in the serial mediation analyses, whereas black pathways indicate non-significant relationships. Significant indirect effects and corresponding 95% confidence intervals are reported for each model.

Significant indirect effects were observed through RAQ scores alone and through the sequential pathway from BKAD to RAQ across all five models. In contrast, indirect effects through BKAD scores alone were not significant for any outcome. Consistent with these findings, RAQ score significantly predicted willingness across all outcomes, whereas BKAD score was not independently associated with willingness after accounting for research attitudes (see Figures 5.1 and 5.2).

## 4. DISCUSSION

The present study investigated factors associated with willingness to participate in Alzheimer’s disease research among a community-based sample of predominantly Black older adults. Specifically, we examined willingness to engage in three research activities related to AD diagnostic procedures, consisting of neuropsychological assessment, blood draw, and brain MRI imaging, as well as two activities related to non-pharmacological interventions to promote brain health, including tracking physical activity via a smartwatch and engaging in aerobic exercise. Research attitudes emerged as the most consistent predictor of willingness to participate across all research activities examined, above and beyond other important characteristics, including age, educational attainment, AD knowledge, family history of dementia, and previous research participation. This finding is consistent with prior research linking positive research attitudes to greater willingness to enroll in AD research studies and procedures (17,27–30). Notably, prior work has identified research attitudes as one of the most consistent predictors of willingness to participate in AD biomarker research across both Black and White participants (29). Given that our sample predominantly consisted of Black older adults, this study did not explore between-group racial differences in willingness to participate in AD research. However, when divided by neighborhood SES level, we found that willingness to participate was broadly similar across levels of neighborhood disadvantage. This finding expands upon prior work by Grill et al., who demonstrated that residents of disadvantaged neighborhoods were similarly willing to participate in research but were less likely to be represented in recruitment registries, suggesting that disparities in research participation may reflect barriers to access and recruitment rather than differences in willingness (15).

Importantly, serial mediation analyses indicated that educational attainment was indirectly associated with willingness through research attitudes and through a sequential pathway involving Alzheimer’s disease knowledge and research attitudes. Higher educational attainment was associated with greater AD knowledge, which in turn was associated with more favorable research attitudes; however, AD knowledge alone did not mediate willingness to participate. The observed association between educational attainment and AD knowledge is consistent with findings reported among older rural adults (31). The present study replicates and extends this association to a predominantly urban, community-based sample.

Crucially, these findings have important implications for optimizing the target and level of interventions to promote research participation. If willingness is comparable across neighborhood contexts, efforts to increase access to research opportunities and reduce barriers to participation may be particularly effective strategies for improving representation among individuals from disadvantaged neighborhoods. A study that featured a culturally tailored AD educational intervention increased both AD knowledge and research participation interest among Hispanic older adults, yet changes in AD knowledge were not independently associated with increased participation interest (20). Similarly, recent evidence demonstrated that trust in researchers was associated with willingness to participate in AD biomarker and neuroimaging research, independent of AD knowledge (9). Another study found that higher educational attainment was associated with both greater AD knowledge and more favorable research attitudes, but only research attitudes were associated with willingness to engage in preclinical AD biomarker testing (32).

Taken together, these findings suggest that AD knowledge alone may be insufficient to promote participation in AD research. Rather, research attitudes appear to represent a more proximal determinant of willingness, serving as a pathway through which knowledge influences participation decisions (9,17,27–29,32). Therefore, interventions that aim to broaden participation in AD research by raising awareness of AD alone are unlikely to increase willingness unless they also address the attitudes and beliefs that ultimately shape participant decisions. Previous studies have documented lower participation among Black adults in Alzheimer’s disease research and have highlighted factors such as trust in researchers, historical injustices, and structural barriers as potential contributors to these disparities (9,17,27–29,32,33). Within this context, our findings further suggest that research attitudes may represent a modifiable target for interventions aimed at improving equitable participation in AD research among predominantly Black community-based populations. Educational campaigns will continue to have value, but they cannot substitute for the long-term relational work of building the conditions under which individuals from underrepresented groups choose to participate.

Overall, participants were generally willing to participate in all study procedures; however, willingness tended to decrease as procedures became more burdensome, with the highest willingness reported for the neuropsychological assessment, followed by the blood draw, and the lowest for undergoing a brain MRI. Neuropsychological assessments are non-invasive and primarily involve completing cognitive tasks, whereas blood draws involve needle insertion and potential discomfort. MRI examinations may present additional challenges, including prolonged immobility and confinement within a noisy scanner environment, potentially contributing to greater apprehension and perceived burden. This pattern highlights how perceived burden, discomfort, or procedural complexity may represent important barriers to participation, a finding that has been most clearly illustrated by the widespread reluctance to undergo AD biomarker detection via lumbar puncture. Recruitment and retention efforts may therefore benefit from strategies that address concerns associated with more burdensome research procedures, such as using a mock MRI scanner.

In contrast to the diagnostic procedures, where willingness appeared to vary primarily according to procedure invasiveness, willingness to participate in the non-pharmacological intervention procedures varied by age. Within this sample of adults aged 50-89 years, older participants reported lower willingness to participate in smartwatch-based research procedures. Older participants may be less familiar with wearable devices, have lower confidence in using new technologies, or be more likely to perceive smartwatches as difficult to operate. Importantly, lower willingness should not be interpreted as evidence that technology-based interventions are unsuitable for older adults. Prior research suggests that technology acceptance among older adults is influenced by perceived usefulness and ease of use, and that wearable devices are more acceptable when they align with user preferences (34,35). Similarly, older participants in our study reported lower willingness to participate in aerobic exercise. This finding should not be interpreted as evidence that older adults are unwilling to engage in exercise-based interventions. Instead, concerns may reflect anticipated physical demands or uncertainty about the ability to participate safely; exercise guidelines emphasize individualized programming and gradual progression based on health status and functional capacity (36). These findings suggest that technology-based interventions should incorporate participant training and user-centered design, while aerobic exercise interventions may benefit from individualized prescriptions and structured ramp-up periods to enhance engagement among older adults.

Our study had several limitations. The present analyses were cross-sectional; therefore, causal relationships between educational attainment, AD knowledge, research attitudes, and willingness to participate cannot be established. Willingness to participate was assessed using responses to specific AD-related research activities rather than a validated measure of overall willingness to participate in research. Additionally, participants in this study had already provided verbal consent as a part of a study exempt from IRB review, which may have resulted in a sample with greater baseline interest in research than the general population. This potential selection bias may limit the generalizability of our findings and could overestimate willingness to participate in AD research. Finally, our outcome reflects self-reported willingness to participate rather than actual research enrollment or participation behavior, and the extent to which these reported attitudes translate into real-world engagement remains unknown.

Future longitudinal and interventional studies are needed to better characterize the directionality and potential causal relationships among education, AD knowledge, research attitudes, and willingness to participate. The development and validation of comprehensive measures of willingness to participate in research, as well as studies examining whether self-reported willingness predicts actual enrollment and participation in research, would strengthen future investigations. Follow-up studies should recruit more representative community-based samples, including individuals with little or no prior research engagement, to better assess willingness across the broader population.

Beyond addressing these methodological considerations, future work should investigate the mechanisms underlying research attitudes and identify modifiable determinants that can improve participation in AD research. In particular, studies should examine whether interventions designed to improve AD knowledge are associated with more favorable research attitudes and ultimately greater participation in AD research. Further characterization of specific dimensions of research attitudes, including trust in researchers, perceived risks and benefits, and concerns about privacy or discrimination, may help identify barriers experienced by different populations. Examining these factors in the context of socioeconomic status, neighborhood characteristics, healthcare access, community context, and other social determinants of health could inform tailored strategies to improve equitable participation and representation in AD research.

## Data Availability

All data produced in the present work are contained in the manuscript

## ACKNOWLEDGEMENTS

This work was supported by a grant from the Alzheimer’s Association (AARFD-24-1309508). The authors used a secure enterprise version of the Gemini generative AI model (Google) accessed via The Ohio State University to generate SPSS syntax for certain statistical analyses and to assist with editing and proofreading the manuscript. All study design, data analysis decisions, verification of statistical results, interpretation of findings, and final manuscript content were reviewed and approved by the authors.

---

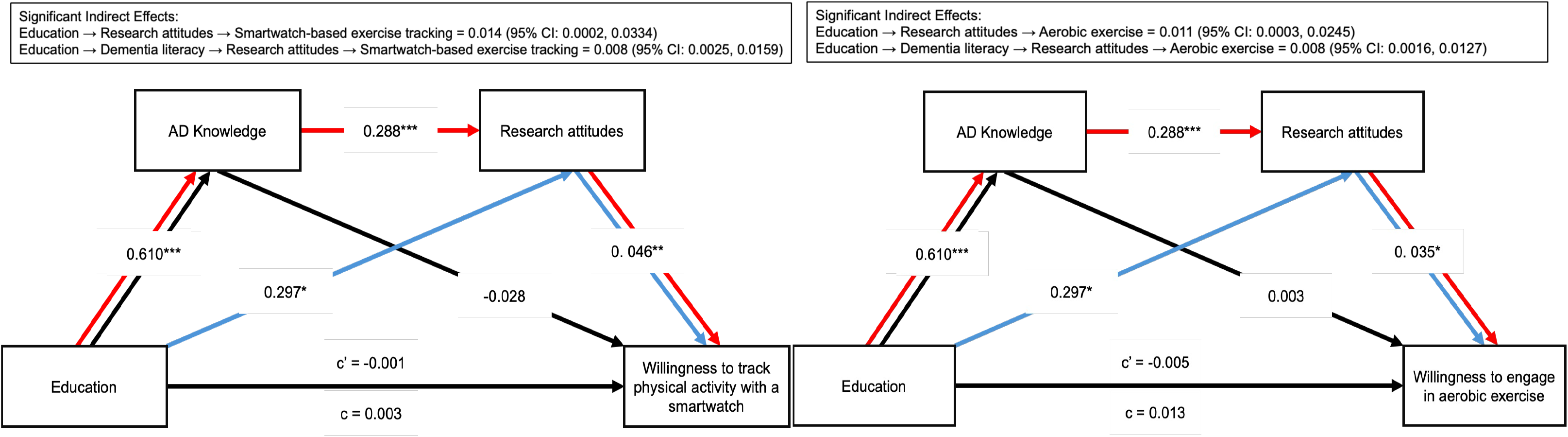

## REFERENCES

1. Gilmore-Bykovskyi AL, Jin Y, Gleason C, Flowers-Benton S, Block LM, Dilworth-Anderson P, et al. Recruitment and retention of underrepresented populations in Alzheimer’s disease research: A systematic review. Alzheimers Dement. 2019;5:751–70. doi:10.1016/j.trci.2019.09.018 PubMed PMID: 31921966; PubMed Central PMCID: PMC6944728.

2. Indorewalla KK, O’Connor MK, Budson AE, Guess (DiTerlizzi) C, Jackson J. Modifiable Barriers for Recruitment and Retention of Older Adults Participants from Underrepresented Minorities in Alzheimer’s Disease Research. J Alzheimer’s Dis. 2021 Apr 6;80(3):927–40. doi:10.3233/JAD-201081

3. Gurland BJ, Wilder DE, Lantigua R, Stern Y, Chen J, Killeffer EH, et al. Rates of dementia in three ethnoracial groups. Int J Geriatr Psychiatry. 1999 Jun;14(6):481–93. PubMed PMID: 10398359.

4. Heyman A, Fillenbaum G, Prosnitz B, Raiford K, Burchett B, Clark C. Estimated Prevalence of Dementia Among Elderly Black and White Community Residents. Arch Neurol. 1991 Jun 1;48(6):594–8. doi:10.1001/archneur.1991.00530180046016

5. Sisco S, Gross AL, Shih RA, Sachs BC, Glymour MM, Bangen KJ, et al. The role of early-life educational quality and literacy in explaining racial disparities in cognition in late life. J Gerontol B Psychol Sci Soc Sci. 2015 Jul;70(4):557–67. doi:10.1093/geronb/gbt133 PubMed Central PMCID: PMC4462668.

6. Kramer M, Cutty M, Knox S, Alekseyenko AV, Mollalo A. Rural–urban disparities of Alzheimer’s disease and related dementias: A scoping review. Alzheimers Dement Transl Res Clin Interv. 2025;11(1):e70047. doi:10.1002/trc2.70047

7. Moreno G, Mangione C, Meza C, Kwon I, Seeman T, Trejo L, et al. Perceptions from Latino and African American Older Adults about Biological Markers in Research. Ethn Dis. 2015;25(3). doi:10.18865/ed.25.3.355

8. Ketchum F, Erickson C, Chin N, Gleason C, Lambrou N, Flowers Benton S, et al. What Influences the Willingness of Blacks and African Americans to Enroll in Preclinical Alzheimer’s Disease Biomarker Research? A Qualitative Vignette Analysis. J Alzheimers Dis. 2022. doi:DOI 10.3233/JAD-215521

9. Raelly-Muze L, Etchison T, Crouch A, Eliacin J, Polsinelli A, Gao S, et al. Association between trust of researchers and willingness to participate in Alzheimer’s disease biomarker research. Alzheimers Dement Behav Socioecon Aging. 2025;1(3):e70024. doi:10.1002/bsa3.70024

10. Williams M, Darcell S, Katherine M, Jonathan H, Pamela J, Morris J, et al. Barriers and Facilitators of African American Participation in Alzheimer Disease Biomarker Research. Alzheimer Dis Assoc Disord. 2010. doi:DOI: 10.1097/WAD.0b013e3181f14a14

11. Shaw AR, Perales-Puchalt J, Johnson E, Espinoza-Kissell P, Acosta-Rullan M, Frederick S, et al. Representation of Racial and Ethnic Minority Populations in Dementia Prevention Trials: A Systematic Review. J Prev Alzheimers Dis. 2022;9(1):113–8. doi:10.14283/jpad.2021.49 PubMed PMID: 35098981; PubMed Central PMCID: PMC8804327.

12. Wendler D, Kington R, Madans J, Van Wye G, Christ-Schmidt H, Pratt LA, et al. Are racial and ethnic minorities less willing to participate in health research? PLoS Med. 2006 Feb;3(2):e19. doi:10.1371/journal.pmed.0030019 PubMed PMID: 16318411; PubMed Central PMCID: PMC1298944.

13. Thetford K, Gillespie TW, Kim Y il, Hansen B, Scarinci IC. Willingness of Latinx and African Americans to Participate in Nontherapeutic Trials: It Depends on Who Runs the Research. Ethn Dis. 31(2):263–72. doi:10.18865/ed.31.2.263 PubMed PMID: 33883867; PubMed Central PMCID: PMC8054877.

14. Eliacin J, Polsinelli AJ, Epperson F, Gao S, Van Heiden S, Westmoreland G, et al. Barriers and facilitators to participating in Alzheimer’s disease biomarker research in black and white older adults. Alzheimers Dement Transl Res Clin Interv. 2023 Jun 5;9(2):e12399. doi:10.1002/trc2.12399 PubMed PMID: 37287470; PubMed Central PMCID: PMC10242196.

15. Grill JD, Kind A, Hoang D, Gillen DL. Diversifying Recruitment Registries: Considering Neighborhood Health Metrics. J Prev Alzheimers Dis. 2022 Jan 1;9(1):119–25. doi:10.14283/jpad.2021.50

16. Witbracht M, Xu Y, Morgan OB, Salazar CR, Hoang D, Kind A, et al. Research Attitudes Questionnaire scores and retention in a recruitment registry. J Alzheimers Dis JAD. 2025 Jan;103(2):406–18. doi:10.1177/13872877241302422 PubMed PMID: 39814522.

17. Stites S. Research attitudes questionnaire scores predict Alzheimer’s disease clinical trial dropout. 2021.

18. Ashford M. Effects of sex, race, ethnicity, and education on online aging research participation. 2020. doi:DOI: 10.1002/trc2.12028

19. Grill J. Retention of Alzheimer’s disease research participants. Alzheimers Dis Assoc Disord. 2019. doi:doi: 10.1097/WAD.0000000000000353

20. Perales-Puchalt J. Preliminary Efficacy of a Recruitment Educational Strategy on Alzheimer’s Disease Knowledge, Research Participation Attitudes, and Enrollment Among Hispanics. Vol. 18. 2019;18(3). 10.1177/1540415319893238

21. Aihara Y, Maeda K. Dementia Literacy and Willingness to Dementia Screening. 2020.

22. Andrade C, Menon V, Ameen S, Kumar Praharaj S. Designing and Conducting Knowledge, Attitude, and Practice Surveys in Psychiatry: Practical Guidance. Indian J Psychol Med. 2020 Aug 27;42(5):478–81. doi:10.1177/0253717620946111 PubMed PMID: 33414597; PubMed Central PMCID: PMC7750837.

23. Wu F, Yang C, Wu F, Sun Z. Knowledge, attitude, and practice toward Alzheimer’s disease prevention among middle-aged and elderly individuals: a cross-sectional study. Dement Geriatr Cogn Disord. 2026 May 16. doi:10.1159/000552507

24. Rubright JD, Cary MS, Karlawish JH, Kim SYH. Measuring how people view biomedical research: Reliability and validity analysis of the Research Attitudes Questionnaire. J Empir Res Hum Res Ethics. 2011 Mar;6(1):1. doi:10.1525/jer.2011.6.1.63 PubMed PMID: 21460589; PubMed Central PMCID: PMC3253733.

25. Wiese LK, Williams CL, Tappen RM, Newman DM. An updated measure for investigating basic knowledge of Alzheimer’s disease in underserved rural settings. Aging Ment Health. 2020 Aug;24(8):8. doi:10.1080/13607863.2019.1584880 PubMed PMID: 30869990; PubMed Central PMCID: PMC8474126.

26. Kind AJH, Buckingham WR. Making Neighborhood-Disadvantage Metrics Accessible — The Neighborhood Atlas. N Engl J Med. 2018 Jun 28;378(26):2456–8. doi:10.1056/NEJMp1802313

27. Erickson CM, Chin NA, Ketchum FB, Jonaitis EM, Zuelsdorff ML, Gleason CE, et al. Predictors of Willingness to Enroll in Hypothetical Alzheimer Disease Biomarker Studies that Disclose Personal Results. Alzheimer Dis Assoc Disord. 2022;36(2):125–32. doi:10.1097/WAD.0000000000000490 PubMed PMID: 35125399; PubMed Central PMCID: PMC9132241.

28. Birnbaum AI, Harkins K, Karlawish J, Grill JD, Gillen DL. Exploring Characteristics Predictive of Willingness to Participate in Preclinical Alzheimer’s Disease Trials. Alzheimers Dement. 2025 Dec 25;21(Suppl 5):e100591. doi:10.1002/alz70859_100591 PubMed PMID: null; PubMed Central PMCID: PMC12740512.

29. Gooding D. Perceptions about preclinical Alzheimer’s disease biomarker collection procedure influences willingness to participate: Findings from an ethnoracially diverse study. 2025.

30. Salazar CR, Hoang D, Gillen DL, Grill JD. Racial and ethnic differences in older adults’ willingness to be contacted about Alzheimer’s disease research participation. Alzheimers Dement Transl Res Clin Interv. 2020;6(1):e12023. doi:10.1002/trc2.12023

31. Wiese LK, Williams CL, Tappen R, Newman D, Rosselli M. Assessment of Basic Knowledge About Alzheimer’s Disease Among Older Rural Residents: A Pilot Test of a New Measure. J Nurs Meas. 2017 Jan 1;25(3):519–48. doi:10.1891/1061-3749.25.3.519

32. Gooding D. Factors Affecting the Willingness of African-American and American Indian/Alaska Native Communities to Engage in Genetic and Biomarker Research: The UBIGR Study. 2024. doi:doi: 10.1016/j.bionps.2024.100090.

33. Zhou Y, Elashoff D, Kremen S, Teng E, Karlawish J, Grill JD. African Americans are less likely to enroll in preclinical Alzheimer’s disease clinical trials. Alzheimers Dement Transl Res Clin Interv. 2017;3(1):57–64. doi:10.1016/j.trci.2016.09.004

34. Ma Q, Chan AHS, Teh PL. Insights into Older Adults’ Technology Acceptance through Meta-Analysis. Int J Human–Computer Interact. 2021 Jul 3;37(11):1049–62. doi:10.1080/10447318.2020.1865005

35. Lassell RKF, Carey CJ, Sample L, Elftmann H, Gitlin LN, Harezlak J, et al. Which activity tracker features matter to you? Older Black participants living with memory challenges and care partner preferences. Innov Aging. 2026 Apr 1;10(4):igag011. doi:10.1093/geroni/igag011

36. Zaleski AL, Taylor BA, Panza GA, Wu Y, Pescatello LS, Thompson PD, et al. Coming of Age: Considerations in the Prescription of Exercise for Older Adults. Methodist DeBakey Cardiovasc J. 2016;12(2):98–104. doi:10.14797/mdcj-12-2-98 PubMed PMID: 27486492; PubMed Central PMCID: PMC4969034.

